# A Versatile Web App for Identifying the Drivers of COVID-19 Epidemics

**DOI:** 10.1101/2020.08.06.20155804

**Authors:** Wayne M Getz, Richard Salter, Ludovica Luisa Vissat, Nir Horvitz

## Abstract

**Background:** No versatile web app exists that allows epidemiologists and managers around the world to comprehensively analyze the impacts of COVID-19 mitigation. The NMB-DASA web app presented here fills this gap.

**Methods:** Our web app uses a model that explicitly identifies susceptible, contact, latent, asymptomatic, symptomatic and recovered classes of individuals, and a parallel set of response classes, subject to lower pathogen-contact rates. The user inputs a CSV file of incidence and, if of interest, mortality rate data. A default set of parameters is available that can be overwritten through input or online entry, and a user-selected subset of these can be fitted to the model using maximum likelihood estimation (MLE). Model fitting and forecasting intervals are specifiable and changes to parameters allow counterfactual and forecasting scenarios. Confidence or credible intervals can be generated using stochastic simulations, based on MLE values, or on an inputted CSV file containing Markov chain Monte Carlo (MCMC) estimates of one or more parameters.

**Findings:** We illustrate the use of our web app in extracting social distancing, social relaxation, surveillance or virulence switching functions (i.e., time varying drivers) from the incidence and mortality rates of COVID-19 epidemics in Israel, South Africa, and England. The Israeli outbreak exhibits four distinct phases: initial outbreak, social distancing, social relaxation, and a second wave mitigation phase. An MCMC projection of this latter phase suggests the Israeli epidemic will continue to produce into late November an average of around 1500 new case per day, unless the population practices social distancing measures at least 5-fold below the level in August, which itself is 4-fold below the level at the start of July. Our analysis of the relatively late South African outbreak that became the world’s fifth largest COVID-19 epidemic in July revealed that the decline through late July and early August was characterised by a social distancing driver operating at more than twice the per-capita applicable-disease-class (pc-adc) rate of the social relaxation driver. Our analysis of the relatively early English outbreak, identified a more than 2-fold improvement in surveillance over the course of the epidemic. It also identified a pc-adc social distancing rate in early August that, though nearly four times the pc-adc social relaxation rate, appeared to barely contain a second wave that would breakout if social distancing was further relaxed.

**Interpretation:** Our web app provides policy makers and health officers who have no epidemiological modelling or computer coding expertise with an invaluable tool for assessing the impacts of different outbreak mitigation policies and measures. To facilitate its use, mouse over instructions, user guides and training videos are available at the website.

## Introduction

The COVID-19 outbreak that in February, 2020, threatened to overwhelm the healthcare system of Wuhan, China, was brought under control in early March through a combination of social distancing, contact tracing and quarantine measures [1, 2, 3, 4]. Since then, these measures have been used by municipalities, counties, states and countries around the world to bring COVID-19 outbreaks under full or partial control. Many of outbreaks have subsequently experienced prominent second waves, as control measures—which we refer to as drivers—have been prematurely relaxed.

As of late August, 2020, the pandemic state of COVID-19 was closing in on twenty five million recorded cases, and over 800,000 recorded deaths. In the US itself, the number of recorded cases was around 6 million and approaching 200,000 recorded deaths. Though the management of outbreaks has been impressive in some countries, in many others healthcare officials and civic leaders at various administrative levels have been in dire need of quantitative tools to help formulate and implement policies designed to flatten and ultimately extinguish state, provincial, and country level epidemics. Currently, policy assessment tools exist only in the hands of computational epidemiologists and modelling and data analysis groups. These individuals and groups are insufficient to meet the policy development and assessment needs of most communities around the world. Without easy entry, versatile, policy evaluation tools, governors, mayors, and other civic leaders responsible for implementing healthcare policy are flying blind when making critical decisions whether or not to open schools or various business sectors representing different levels of SARS-CoV-2 transmission risk.

Here we present an easy entry, user-friendly analytical tool: the COVID-19 NMB-DASA (Numerus Model Builder Data and Simulation Analysis) web app. It can be used to address questions regarding the impact of social distancing, social relaxation, changes in surveillance, implementation of contact tracing with quarantining, patient isolation, and vaccination (when widely available) on incidence and mortality rates. NMB-DASA does not require the user to have a mathematical or epidemiological modelling background or an understanding of the computational procedures needed to carry out deterministic and stochastic simulations for model parameter estimation or epidemic forecasting. It only requires users to provide a comma separated values file (CSV: a standard used by all common spreadsheet applications and data management programs) that contains the incidence and mortality time series of the particular outbreak to be analysed, to read this paper, and to undergo less then an hour of training using videos supplied for this purpose at the website. The user is also able to load a set of generic default parameters that come with our COVID-19 NMB-DASA, or modify these by either entering new values or manipulating sliders on web pages. We also illustrate, using data from the Israeli, South African, and English COVID-19 epidemics, how the NMB-DASA web app can be used to: i) forensically analyze the dynamic aspects of outbreaks by identifying implicit drivers (Viz., surveillance levels, social distancing and relaxation, quarantining, improvement in therapeutics, rolling out vaccination programs); and, ii) evaluate or forecast the impacts of changes to these drivers.

## Methods

### Model

The epidemiological simulation and forecasting engine of our website is based on a modified SEIR (<u>S</u>usceptible, <u>E</u>xposed, <u>I</u>nfectious, <u>R</u>ecovered) formulation called SCLAIV (Fig. 1), which expands disease class E into a contact class C and latent class L. Individuals in C, having made contact with SARS-CoV-2, can either thwart potential infection (e.g., through physical barriers or operation of the innate immune system) and return to S or succumb and moving onto L. SCLAIV also adds an asymptomatic infectious class A to the general infectious component of an SEIR process, which is known to be important component of SARS-CoV-2 transmission [5, 6, 7, 8]. Individuals in A are assumed to be less infectious than in the class I [9] and can either move to I or directly to the acquired immunity (i.e., akin to naturally “vaccinated”) class V. A SCLAIV+D formulation also explicitly separates out the removed class R in the SEIR process into V and the class D of individuals that have died.

**Figure 1.**
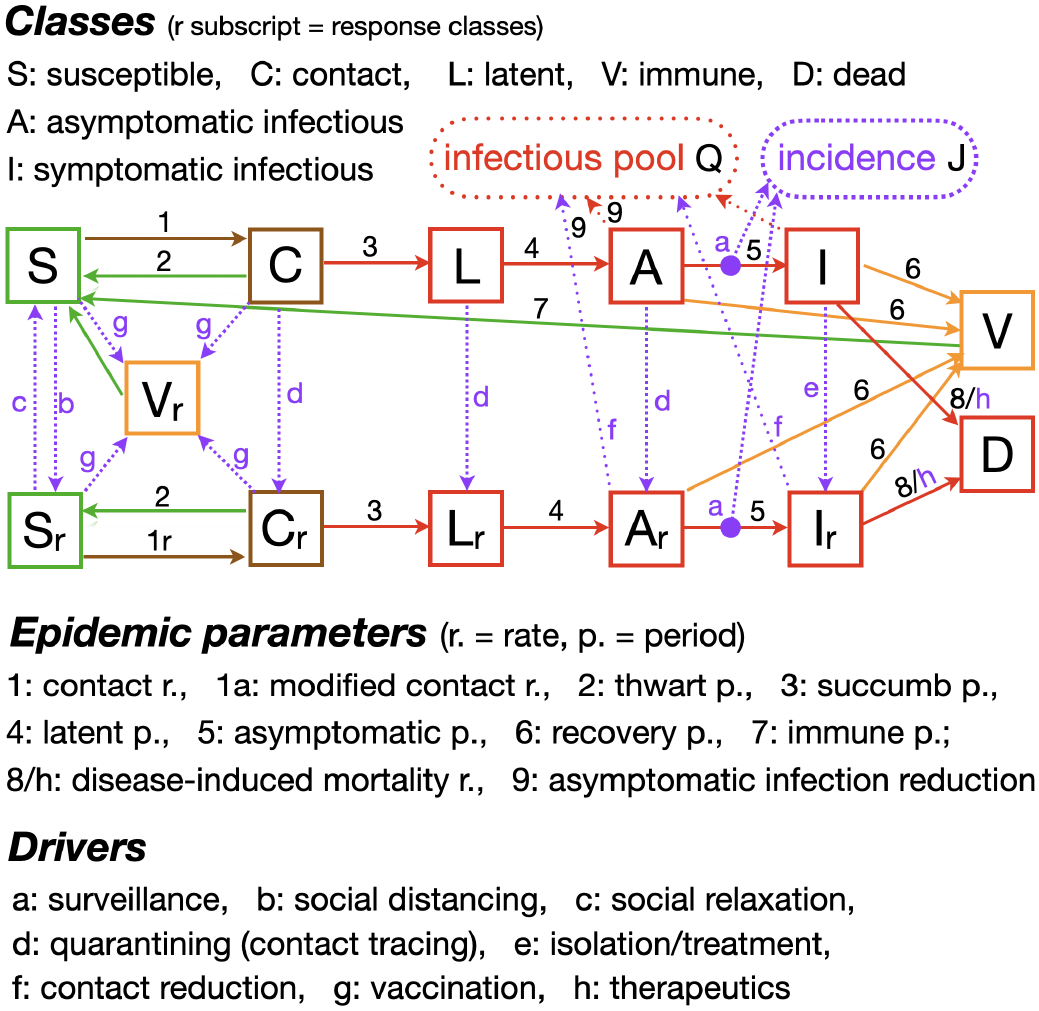
A flow diagram of the SCLAIV+D+response model with its 8 flow parameters identified by Roman numbers 1-8 plus an asymptomatic infection parameter 9. The 8 drivers identified by lower case Roman letters a-h are either zero (apart from surveillance), a positive constant or have the form of a switching function, as described in the text. We have generalized the disease-induced natural mortality flow rate 8 from being a constant to include the possibility of driving (h) the rate down to account for improvements over time in treatment and therapeutics.

SEIR and SCLAIV+D formulations need to be augmented to include the epidemiological driving process used to “flatten incidence and mortality curves” during outbreaks, if models are used to assess the impacts of healthcare policies on epidemics. The structure and equation details of our full SCLAIV+D+response model are provided in Appendix **A** of the Supplementary Material (SM). Details of our discrete time deterministic and stochastic implementations of our SCLAIV+D+response model, at the computational core of our web app, can be found in Appendix **B** (SM). This implementation includes 8 response drivers (Fig. 1), each of which can either be specified as a constant or time varying rate—the latter, specifically, as a five parameter switching function (Appendix **A.3**; Fig. A.2. SM). These driving rates are responsible for transferring individuals among basic SCLAIV and SCLAIV-response classes (Fig. 1). Specifically, they are a social distancing rate (transfers individuals from S to S_r_), which involves a possible dynamic change in a contact rate reduction parameter, a social relaxation rate (transfers individuals from S_r_ to S), a quarantine rate linked to contact tracing (transfers individuals from C, L and A to C_r_, L_r_, and A_r_ respectively), case isolation (transfers individuals from I to I_r_) and vaccination rates (transfers individuals from pre-infectious classes to V_r_; see Fig. 1), the level of surveillance (i.e., proportion of cases detected) implemented to monitor the state of the outbreak, and the impacts of treatment on disease-induced mortality rates (the latter is also referred to as the virulence).

### Parameters and data

Many, but not all, of the basic epidemiological parameters used to model COVID-19 and other infectious disease outbreaks can be obtained independently from data on the case histories individuals infected with SARS-CoV-2 [16, 17, 3, 10], while others can be estimated by fitting models to incidence, mortality and other types of data. Those estimated prior to model fitting (Table 1) are likely to include the latent, asymptomatic, recovery (i.e., from classes A, A_r_, I and I_r_ to V), and immune periods (the latter can only be estimated from future data, depending on the mean period of acquired immunity that will be realised over time), because these are more directly observable from clinical studies than, for example, contact rates and transmission probabilities associated with particular types of contact events. We also note that many parameters are correlated because they act together to produce particular types of observable phenomena, such as growth rates or changes in incidence or mortality levels from one point in time to the next. Thus, for example, contact rates, probability of pathogen transmission per contact, and average infectious period length are linked when it comes to generating the more directly estimable quantity called *R*_0_ [18], which is the average number of individuals an infected individual can expect to infect at the initial stage of an outbreak. So, if a value for the infectious period is selected a *priori* to be higher than it really should be, fitting a contact rate parameter value to the incidence time series using, say, maximum likelihood estimation (MLE) will yield a concomitantly lower value for this rate than if the *a priori* infectious period had been lower.

In Table 1 a default set of point estimates of parameter values are listed for modelling the initial phase of a non-specific COVID-19 outbreak (also see Fig. 2). These values are used to derive probabilities of disease progression events at the individual level, scaled up to the population level using multinomial distributions (stochastic simulations) or proportions of individuals undergoing disease class transitions (deterministic simulations) (see Appendix **B** of SM). Further, these values can be overridden when values more appropriate to a specific region are available, or values can be selected according to their likelihoods from a distribution of values when available from a suitable source (e.g., from an MCMC procedure, as discussed in Appendix **C**, SM). Of course, incidence and mortality time series data, as well as response driver parameters, will be region specific and depend on policies implemented to help control local outbreaks.

**Table 1.** Default point estimates of parameters for fitting and simulating a basic COVID-19 SCLAIV outbreak on [0, *t*_est_]). Also see Fig. 2

| Parameter | Symbol(math<sup>†</sup>) | Value | Source/Comment |
| --- | --- | --- | --- |
| **Transmission** |  |  |  |
| Contact rate<sup>§</sup> | $\kappa$ | estimated | incidence data* |
| Nominal/effective pop size | $N_0 = N_{\text{eff}}$ | $10^5 - 10^7$ | varies<sup>¶</sup> |
| Asymptomatic reduction | $\varepsilon$ | 0.1 | unknown: see [10] |
| Surveillance | $\delta_{\text{sur}}$ | 0.5 | unknown** |
| Isolation/treatment | $\delta_{\text{iso}}$ | 0.35 | varies*<sup>††</sup> |
| Contact rate reduction | $\delta_{\text{con}}$ | 0.1 | varies*<sup>‡</sup> |
| **Progression** |  |  |  |
| Thwart period | $\pi_{\text{thw}}$ | 1 | normalised |
| Succumb period<sup>§</sup> | $\pi_{\text{suc}}$ | estimated | incidence data* |
| Latent Period | $\pi_{\text{lat}}$ | 4<sup>††</sup> days | [7, 11, 10] |
| Asymptomatic period | $\pi_{\text{asy}}$ | 5<sup>††</sup> days | [7, 11, 10] |
| Symptomatic/recovery period | $\pi_{\text{rec}}$ | 7<sup>‡</sup> days | [10] |
| Immune period | $\pi_{\text{imm}}$ | 1000 days | unknown |
| Disease-induced mort. | $\alpha$ | estimated | mortality data |
| **Initial Values** |  |  |  |
| Initial Susceptible | $S_0$ | $S_0 = N_{\text{eff}} - C_0 - L_0 - A_0 - I_0 - V_0$ | |
| Initial Contact | $C_0$ | estimated | incidence data* |
| Initial Latent | $L_0$ | $\left(\frac{\pi_{\text{asy}}}{\pi_{\text{rec}}}\right) G I_0$ | requires $G^+$ |
| Initial Asymptomatic | $A_0$ | $\left(\frac{\pi_{\text{lat}} \pi_{\text{asy}}}{\pi_{\text{rec}}^2}\right) G^2 I_0$ | requires $G^+$ |
| Initial Symptomatic | $I_0$ | estimated | incidence data* |
| Initial Immune | $V_0 = 0$ | SARS-CoV-2 immunologically naïve pop. | |

<sup>§</sup>See Eq. B.3 in Appendix B of SM for relationship to SEIR model parameter $\beta$
<sup>†</sup>The equivalent symbols used by the web app are $\kappa = \text{kappa}$ , $\pi_{\text{thw}} = \text{P.thw}$ , $I_0 = \text{I}_0$ , etc., as will be made clear throughout the text
\*Best values estimated using likelihood optimisation, ranges estimated using Bayesian MCMC methods [12, 13, 14]
<sup>¶</sup>Depends on relative outbreak size (e.g., < 10% of total pop.) for reliable forecasts
\*<sup>††</sup>Varies. This default value implies individuals will be isolated within three days of entering the symptomatic class
\*<sup>‡</sup>Varies. This rate assumes that sheltering-in-place reduces contact rates by 90%
\*\*Values fitted to the initial conditions will inversely scale with this value
<sup>†</sup>Sources are highly variable, so we selected integer-valued ball park estimates
<sup>‡</sup>Value early into epidemic since it depends on hospitalisation/isolation protocols
<sup>||</sup>If sufficiently large, its influence on the initial stage of the outbreak is negligible
<sup>+</sup>Requires $G$ which, unlike $R_0$ , is a rate of increase over the recovery rather than the “serial” interval [15, 1]

The disease induced mortality rate can also be obtained directly from epidemiological data and the average mortality rate over any period of time can be estimated from the number cases observed and the ultimate fate of those cases: recovery or death. During the course of an outbreak, in the context of COVID-19, however, the time between an individual being infected with SARS-CoV-2 and a definitive outcome can be several weeks. So this time lag has to be taken into account when estimating mortality [10]. Further, during the course of an outbreak, provided the healthcare system is not overwhelmed, we might expect mortality rates to drop as hospitals learn how to take care of the more serious cases admitted for treatment and care.

**Figure 2.**
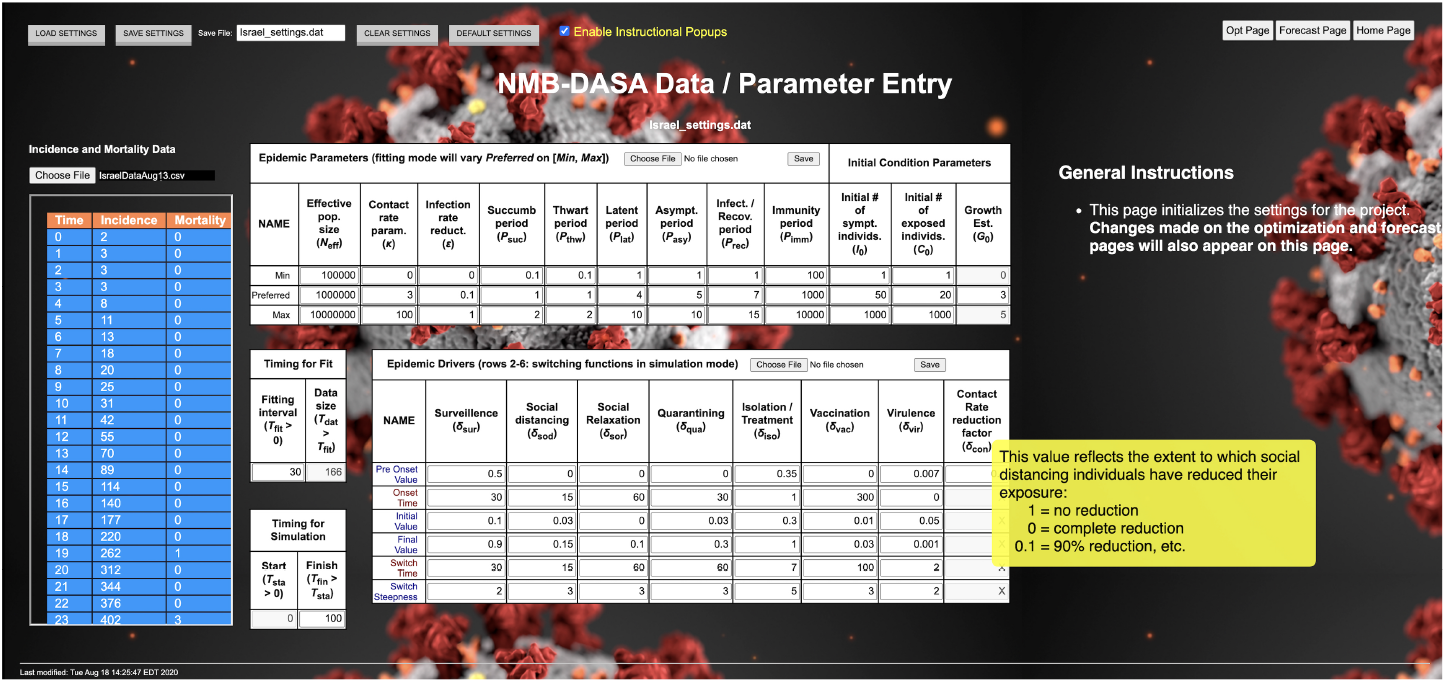
Incidence and Mortality time series are loaded by selecting a CSV file (see the Choose File window on the left). These data along with Epidemic Parameters and Epidemic Drivers values can be loaded separately or default values (shown here) can be loaded, modified, and then saved for use later as a single.dat file. The Fitting interval value is inserted by the user and the Data size value is automatically computed from the length of Incidence and Mortality time series. Once all the parameter values have been set, the Opt Page button on the right can be selected to take the user to the NMB-DASA optimisation page. A forecasting simulation Finish time can be inserted or selected later on the Forecast Page. The yellow Enable Instructional Popups window can be checked to activate mouse-over popups, as illustrated here for the Contact Rate reduction factor.

Parameters that are difficult to explicitly identify from case-history data include the absolute contact rates as a function of behaviour (social distancing or not) and the probability of transmission as a function of the proximity, duration, and background environmental conditions (e.g., humidity, temperature, air movement, fomite characteristics [19] during contact). Also difficult to obtain is the state of the population at the time that the first few cases are detected. The first cases are the first group of individuals identified in disease class I at the nominal start of modelling an outbreak, which for convenience we denote as time *t* = 0. Of course, we anchor this point to some suitable calendar day to facilitate interpretation of results. The number of individuals assigned the value *I*(0) depends on the level of surveillance at time t. In a well-developed health care system, depending on the proportion of asymptomatic carriers and the severity of the disease, more than half the cases may easily go undetected during the first few weeks of an outbreak, though surveillance should improve during the course of the outbreak. In a poorly-developed healthcare system, the proportion of cases detected during the initial stage of an outbreak may be considerably lower.

Of course, only symptomatic cases will initially be detected: asymptomatic cases will require the implementation of some sort of testing regimen. Thus, at the start of an epidemic, the numbers of individuals in disease classes C (*C*(0)), L (*L*(0)) and A (*A*(0)) are unknown and, thus, need to be estimated. The latter two can be expressed in terms of the estimated value *I*(0) if the initial growth rate of the epidemic can be reasonably guessed at from past experience with similar epidemics, in which case *L*(0) and *A*(0) may computed from the estimate of *I*(0) (see Appendix B.2, SM) rather than having to be estimated as well. The value of *C*(0), however, also needs to be estimate since its relationship to *I*(0) depends on various other parameters, such as the thwart and succumb periods. Since these two periods are complementary rates out of the same disease class C, and (as a first cut) the net rate through C is more critical to estimate than the size of C itself, we can fix the thwart period at 1. In this case, the fit of the contact rate and succumb periods will then scale relative to the thwart period. The sensitivity of results to this first-cut assumption can later be tested or relaxed, as deemed appropriate.

The initial value *V* (0) for the number of individuals assumed to be immune to infection by SARS-CoV-2 at the start of the outbreak can be set to be 0, since this virus is thought to be novel for humans. Of course, it may emerge that some cross immunity exists for individuals that have previously been exposed to SARS-CoV1 or other corona viruses [20]. In this case, initial immunity data can be used to estimate a value for *V* (0) or we can think of the population at risk, as discussed below, to be some appropriate fraction of its true size. In this case, a slightly higher estimate of the true contact rate will emerge from fitting the model to incidence data because contact rates are inversely proportional to the population size *N*(*t*) (see Eqns. A.17 in SM).

Initially, two of the drivers—surveillance and patient isolation rates—can be set at positive constant values, based on a crude expectation of the current level of efficiency of the healthcare infrastructure. For a developed country, we may want to set initial surveillance levels at, say, 0.5 (50%) and hospitalisation levels at, say, 0.35, or some other values that are more reflective of the severity or mildness of the disease. For countries with poor health infrastructure, or diseases milder than COVID-19, values of 0.1 for both these levels may be more suitable. These guesses can be changed later, as the forensic analysis of the structure of the outbreak proceeds, and the sensitivity of projected outcomes to these initial guesses are evaluated.

Another parameter that remains somewhat uncertain at the start of an outbreak is the size *N* of the population at risk. In an isolated, spatially homogeneous population (e.g., an island where all individuals randomly encounter others from different parts of the island—that is, the population is well mixed at, say, a weekly scale), *N* should be the size of the population. In large metropolitan areas where the boundaries separating the relatively well-mixed portion of the population compared to outlying areas, we may want to set the value of *N* to some nominal/effective value *N*_eff_ that is, say, 1 million or even 10 million when the population size justifies this choice. Initially the actual choice is not critical, because the outbreak dynamics in standard SEIR models, including SCLAIV, is insensitive to the value of *N*_eff_ [21]. As the outbreak progresses and the total number of infections approaches, say 10% of *N*_eff_, then our choice for this value becomes significant. The reason is that *N*_eff_ determines how rapidly the exponential phase of the initial outbreak begins to lose steam as the decreasing proportion of susceptible individuals in the population begins to play a role in cresting the incidence curve. Thus for epidemics in which the total number of infections exceeds 10%, it may be useful to estimate the “effective population size” *N*_eff_ when fitting the model output to incidence data [21].

### Fitting the model

**Figure 3.**
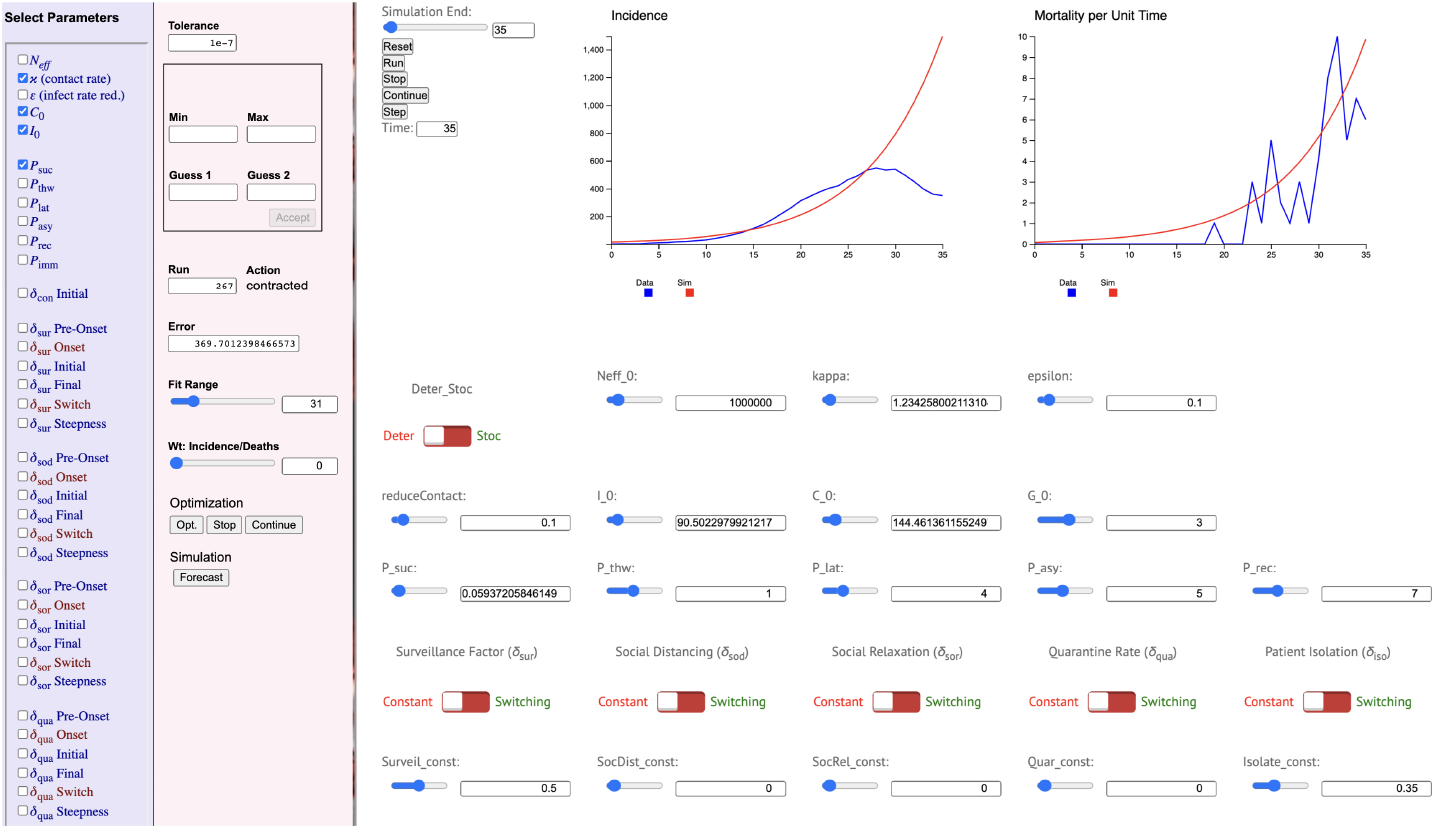
An incomplete view (upper left corner) of the optimisation page of the NMB-DASA web app. The leftmost (purple) panel is where variables to be included in the optimisation are checked. The next (pin) panel to its right is where the details of the optimisation procedure are entered: in this case the Fit Range is 31 and Wt:Incidence/Deaths is zero, implying that only the incidence curve is being fit. In this pink panel we see the MLE took 267 iterations to converge, and the absolute log-likelihood error (369 after truncating beyond the decimal point; see Eq. C.2, SM) is provided in the Error window. The incidence and mortality time series and simulated solutions over the interval [0,35] are plotted in top part of the white panel. Below these are MLE and input values of the basic SCLAIV parameters used in the simulation, apart from the mortality rate (i.e., virulence: for this fit has been set to 0.001) that on a larger view of the Opt Page appears just to right of the Patient Isolation window. All switching functions are set to the Constant mode (red rather than green switch values), with indicated values in this case provided rather than fitted.

NMB-DASA uses a maximum-likelihood estimation (MLE) method to fit selected parameters to the incidence and simultaneously or separately to the mortality data by selecting a value for `Wt:Incidence/Deaths`, as discussed below (also see Fig. 3). It also provides for parameter values obtained from Bayesian Markov chain Monte Carlo (MCMC) methods [13, 22] generated elsewhere to be used in forecasting simulations (See Section C.3, SM). We plan to include MCMC directly in future versions of NMB-DASA

The following is an outline of the process we used to undertake a forensic analysis of the Israeli incidence and mortality rate data and, in part, the South African and English incidence and mortality rate data. In all cases, we fitted our model to a 7-day lagged moving average of the incidence data because of an obvious seven-day oscillation in new cases reported around the world [23] and, also, newly identified cases are likely associated with individuals actually transferring to the I class some days prior to this reporting event.

A step-wise procedure for running and fitting the SCLAIV model (also see the videos provided at the website) is:

1. *Load Data*. Load the incidence and mortality data using a CSV (comma separated values) file in which the first row is an incidence data time series and the second row is a mortality time series of the same length as the first. Load the default set of epidemic and driver parameters. If any of the default parameters require modification, these can be overwritten by directly entering data into the appropriate window and then saved by selecting the SAVE SETTINGS button for direct loading next time around.
2. *Fit Initial Outbreak Phase*. Fit those basic SCLAIV epidemic parameters that cannot be independently estimated by checking them off on the optimisation page (purple panel in Fig. 3), setting `Wt:incidence/deaths` to 1, `Fit Range` to some desired value, such as 14 or 30, and then pressing the `Opt` button (pink panel in Fig. 3). These parameters are likely to be the contact rate `kappa`, the succumb period `P_suc`, the initial value `C_O` for the contact class, and the initial value `I_0` for the symptomatic infectious class. Note, as discussed above, the ratio `P_suc/P_thw` is more salient than either parameter on its own, so we have normalised `P_thw=1`. After running an optimisation, the MLE values obtained can be saved using the SAVE SETTINGS button. If desired, an MLE value for the mortality rate can be obtained by unchecking all the variables use in the previous fitting, checking `vir_const`, resetting `Wt:incidence/deaths`=0 and then pressing the `Opt` button again. We note that fitting incidence and mortality are not entirely independent processes, but they are nearly so while the proportion dying is just a few percent and the proportion of susceptibles remains greater than 90%. By selecting `Wt:incidence/deaths` in between 0 and 1 we can fit both simultaneously, but are then faced with obtaining different solutions for different values of the `Wt:incidence/deaths` setting on (0, 1).
3. *Fit Social Distancing Phase*. When using the model to project the outbreak pattern beyond the initial fit phase, a point will likely be reached where the simulated incidence time series is growing much more rapidly than the empirical time series. If the reverse occurs, then the *Fit Initial Outbreak Phase* step described above should be repeated using a shorter `Fit Range` value. The reason for this over estimation is either because the effects of social distancing are beginning to take affect, or the effective population size is considerably smaller than initially anticipated. If the latter case is suspected then *N*_eff_ should be increased, or it can be replaced by an *N*_eff_ parameter whose size is estimated as part of the MLE process, as discussed in [21]. At this point, one can proceed by unchecking all variables fitted in the *Fit Initial outbreak Phase* optimisation procedure, and then checking all the social relaxation driver variables to carry out a new MLE fitting procedure with the `Fit Range` parameter now set beyond its *Fit Initial Outbreak Phase* by several weeks, using the data as a rough guide to where the end of the social distancing phase appears to be occurring because incidence is beginning to decline less steeply or even rise (e.g., see Fig. 4).
4. *Fit Social Relaxation Phase*. The model can now be used to project the outbreak pattern beyond the *Fit Social Distancing Phase*. If the outbreak has been successfully quelled then the fitting task is complete. In many cases, however, the outbreak may either not be dampened sufficiently fast (as in the case of the COVID-19 in England, see Section 3.3 for details) or may even exhibit a second growth phase that is stronger than the first (Fig. 4). In either case, the MLE values obtained in the *Fit Initial Outbreak Phase* and *Fit Social Distancing Phase* can be saved and a third round of MLE fitting executed. In this case we may choose to check only the social distancing driver parameters and optimise over a `Fit Range` that goes several weeks beyond the *Fit Social Distancing Phase*. Other approaches can be taken, discussed in the different case studies below.
5. *Scenario Projection and Counterfactual Assessments*. We have considerable freedom to run counter-factuals and forecasts (see Appendix C.2, SM), depending on what values we give to the `Fitting Interval` and `Finish Timing for simulation` variables (Fig. 2). Forecasting using MCMC parameter estimates can also be undertaken (see Appendix C.3, SM), as illustrated in the Israel COVID-19 case study. The forecasting page of the web app is designed to permit the flexibility of working with two data sets and two time windows: the first is to simulate the model in deterministic mode on an interval [0*,t*_fit_] and second to allow one (in deterministic mode) or repeated runs (in stochastic mode) on the interval [*t*_fit_*,t*_fin_]. The latter can be either executed by incorporating demographic stochasticity alone, or it can include a file of parameter values obtained from an MCMC fit of the model to data.

**Figure 4.**
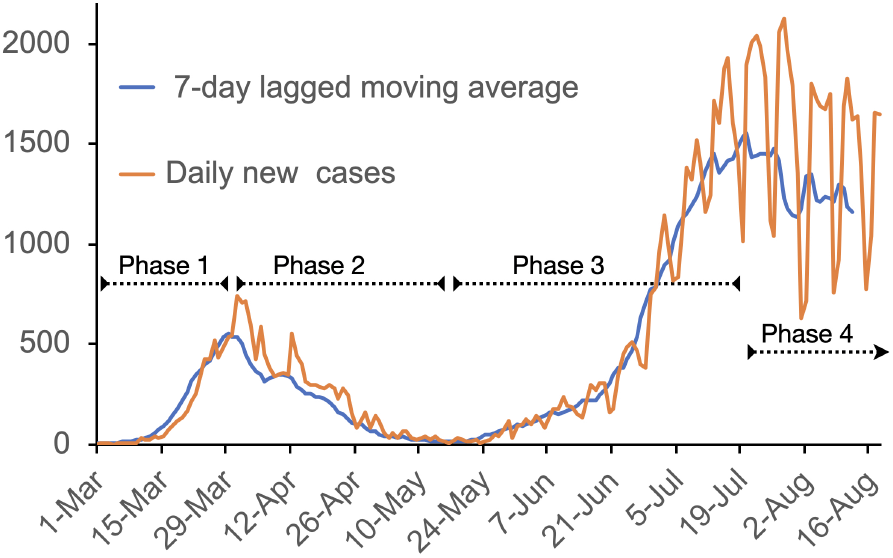
The daily incidence (new cases) time series (points connected by light brown lines) is plotted here from March 1, 2020 to August, 18, 2020. The blue curve is a 7-day lagged moving average plotted to August 12 (which averages over data up to August 18: deaths on August 12, for example can only be identified and recorded as caused by COVID-19 on or after the actual day of death). The different phases of the outbreak, approximately indicated by broken range bars, are: 1. *Initial outbreak phase*, 2. *Social distancing phase*, 3. *Social relaxation phase*, 4. *Second wave mitigation phase*.

## Illustrative Studies

The Israeli, South African, and English COVID-19 studies presented here are of limited scope. Deeper studies typically involve teams of individuals including those familiar with any idiosyncrasies of the data. Thus our examples are meant to be purely illustrative of how NMB-DASA can be used rather than comprehensive analyses of the epidemics considered.

### COVID-19 in Israel

**Figure 5.**
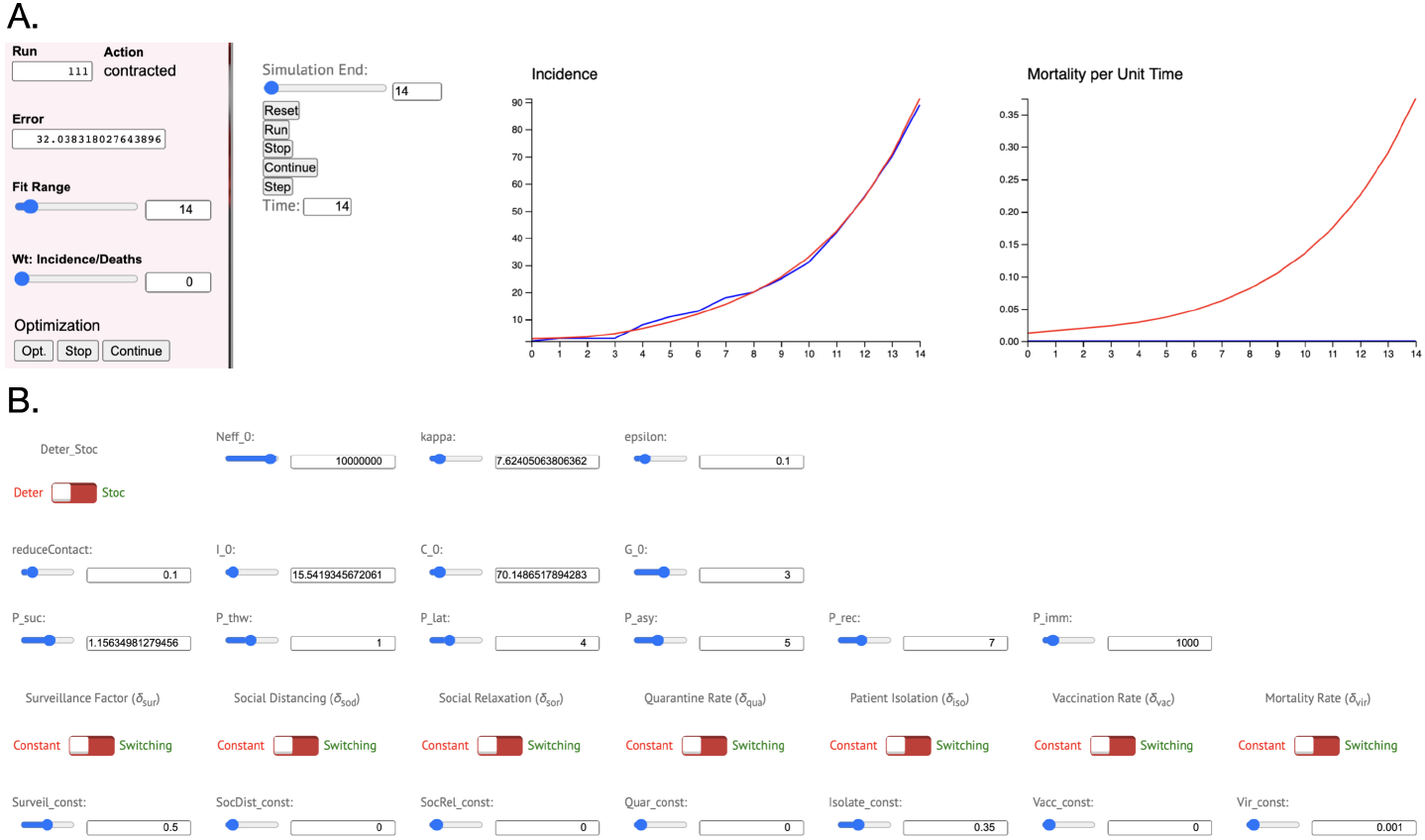
A. MLE of the basic SCLAIV parameters fitted to the first 14 days (March 1 to 14) of Israeli incidence data only (Wt:Incidence/Deaths=0). B. MLE values obtained from this fit. Note, the mortality simulation does not rise to a single individual since no COVID-19 deaths were recorded in Israel during the first two weeks of March. Compare this fit with the same fitting procedure applied to the whole of March, as depicted in Fig. 3, where we see a rising daily death total in the second half of March.

The first COVID-19 infections in Israel occurred in late February [24, 25]. A steady stream of cases only began to be recorded starting around March 1. On March 9, a 14-day home isolation was imposed on all individuals arriving from abroad. A national state of emergency was declared in the Israel on March 19 with the first recorded COVID-19 death of an Israeli citizen occurring on March 20 [24, 25].

A plot of the COVID-19 incidence data from March 1 to August 18 (7-day lagged moving average to August 12) in Israel reveals a rising *initial outbreak phase* in March, followed by a steady drop during a *social distancing phase* to an average of fewer than 20 new cases per day during the second and third weeks of May (Fig. 4). During the last week of May, incidence began to rise steadily again during a *social relaxation phase*. A *second wave mitigation phase* set in around mid-July, as the second wave was flattened in response to measures used to try and bring the epidemic under control [26].

We begin by first using MLE to fit our basic free set of SCLAIV parameters (i.e., values for the contact rate `kappa`, succumb period `P_suc`, and initial number of individuals in classes C (`C_0`) and I (`I_0`), which were not set *a priori* from literature data—see Table 1) to the first two weeks of the *initial outbreak* data in March 2020. The fit was tight, as shown in Fig. 5 along with the four MLE values (Fig. 5 obtained from the optimisation procedure and other relevant SCLAIV parameters used in model. A subsequent fit to the first month of data indicated that the effects of social distancing were already retarding the outbreak in the second half of March (Fig. 3), yielding a new set of four MLE values applicable to fitting the model over the first 31 days. This 31-day fit, however, was quite poor, so we decided to added a social distancing component to MLE fitting of the *initial outbreak* phase (Fig. 6A).

**Figure 6.**
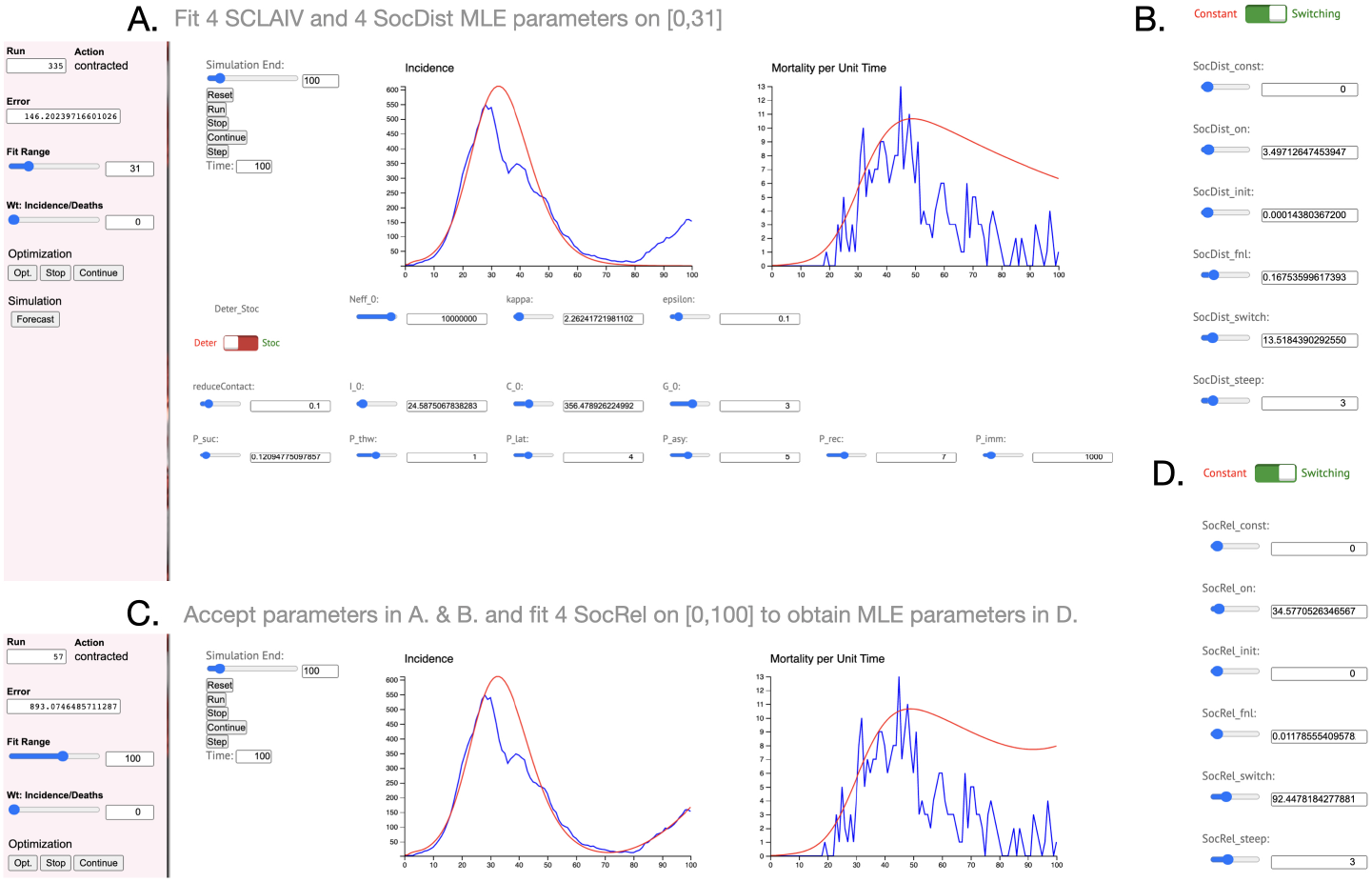
A & B.. An MLE _t of the basic SCLAIV model & 4 social distancing parameters (SocDist steep fixed at 3) to the first 31 days of incidence data. C. & D. An extension of the fit in A obtained by accepting the MLE parameters listed in A & B but fitting 4 social relaxation parameters (SocRel steep fixed at 3) to the first 100. Note the value SocRel init=0 in D was not fixed, but represents a boundary constraint in the MLE solution.

We thus added a social distancing switching function to our MLE fit of incidence over the first 31 days to obtain Fig. 6A, which includes simulation of the model out to day 100. This simulation indicates that the 31-day MLE continues to fit incidence well to around day 80, but then fails to fit to day 100. To correct this, we could either: i.) fix the 8 MLE parameters obtained in our 31-day fit (`kappa, I_0, C_0, P_suc` in panel Fig. 6A and `SocDist_on, SocDist_init, SocDist_fnl, SocDist_switch` in panel Fig. 6B), but now check the 4 social relaxation parameters `SocRel_on, SocRel_init, SocRel_fnl, SocRel_switch` in panel Fig. 6D to obtain a new 4 parameter MLE fit over the first 100 days; or ii) we could simultaneously fit all 12 parameters to the first 100 days. We followed the former case to obtained the fit illustrated in Fig. 6C and the 4 new MLE parameter values provided in Fig. 6D. Again we note that `SocRel_init`=0 is part of a solution induced by the constraint `SocRel_init`≥ 0.

We also decided to see how well our MLE procedure would perform when fitting all 12 parameters simultaneously, but we did this in the context of all 135 days of incidence data. The result of this MLE undertaking is illustrated in Fig. 7A. In both of the above fits to the first 100 and then 135 incidence data points, we set the disease-induced mortality rate to 0.001 (i.e., `Vir_init`=0.001). The fits of the mortality rate time series thereby obtained, appear reasonable for the initial phase of the outbreak, but fail beyond the third week of April (e.g., see middle graph in Fig. 7). We thus decided to fit a switching curve to the mortality data, by fixing all the values used to obtain the MLE fit depicted in Fig. 7A, setting `Wt:Incidence/Deaths` to 1, and allowing the parameters `Vir_init`, `Vir_fnl`, and `Vir_switch` to vary in an MLE fit of the model to the mortality data. The resulting MLE fit is depicted in Fig. 7B. We note that the values for `Vir_onset` and `Vir_steep` were *a priori* set at 0 and 1 respectively, since disease-induced mortality is always in effect. The remaining MLE parameter values for the virulence switching curve are listed in Fig. 7B. Again we note that the MLE value `Vir_fnl`=1 represents a boundary solution. A plot of the three switching functions associated with the fits depicted in Fig. 7 are provided in Fig. 8.

**Figure 7.**
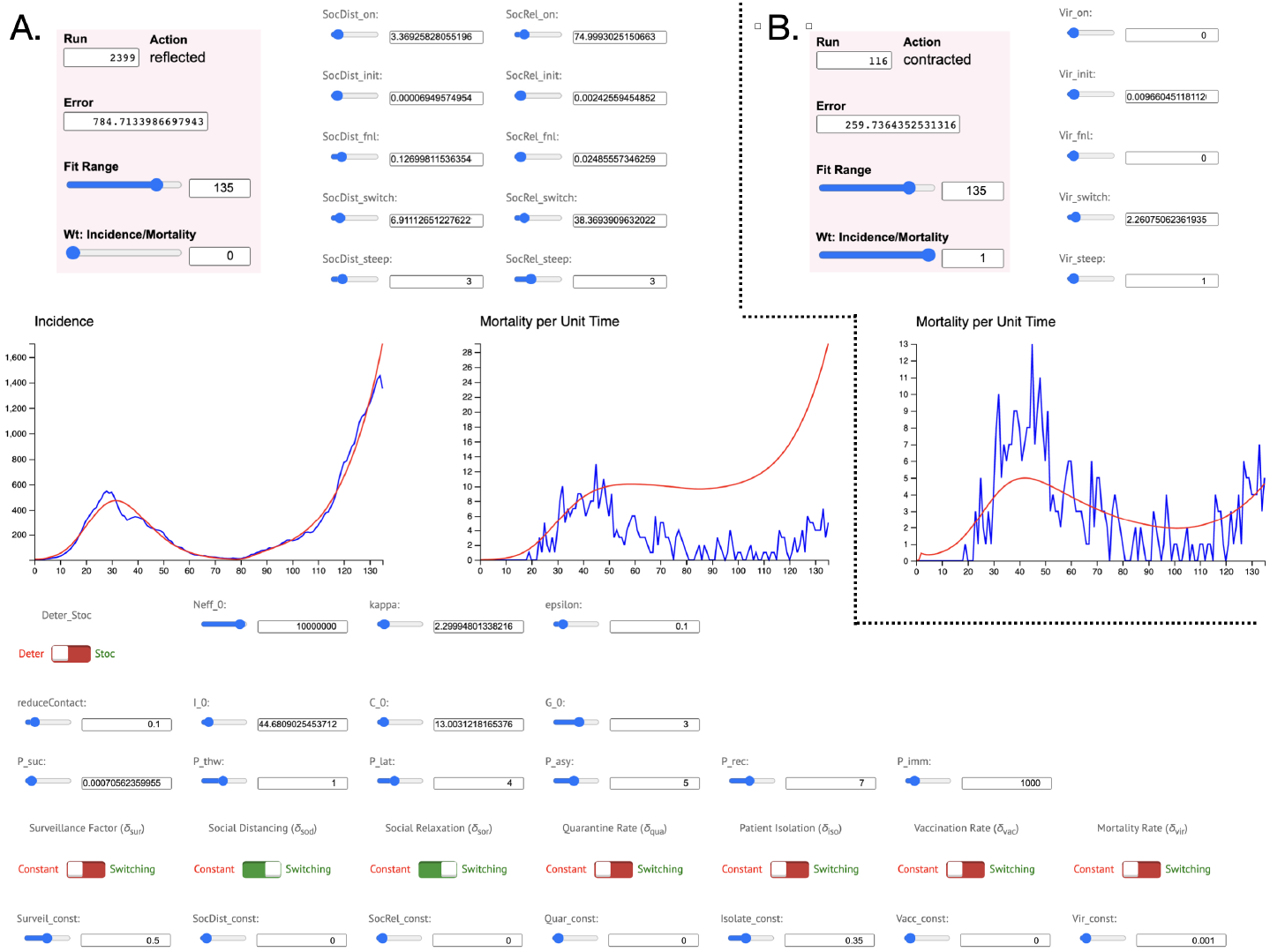
A. Simultaneous MLE of 4 SCLAIV, 4 social-distancing and 4 social-relaxation parameters to the first 135 incidence time series points (i.e. Wt:Incidence/Deaths=0). B. A better fit of the mortality data alone can be obtained by accepting the MLE values in A, setting Wt:Incidence/Deaths=1 and obtaining ML fit for the 3 virulence parameters (we fixed Vir on=0 and Vir steep=1) to the first 135 mortality time series points. Note that Vir_fnl=0 is a boundary constraint solution.

**Figure 8.**
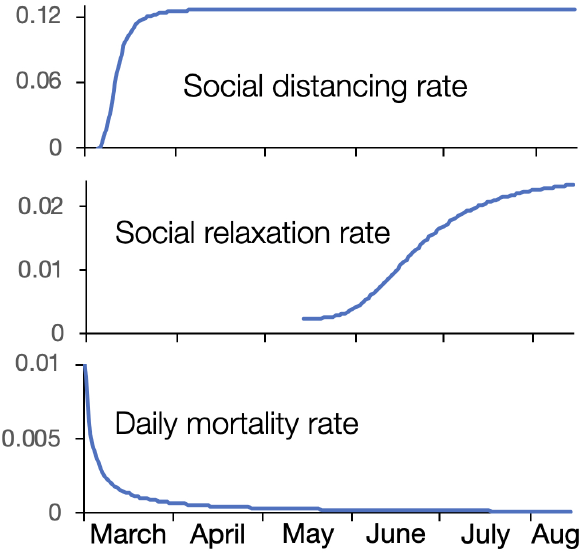
Plots of the social-distancing switching (applied only to class S_r_), social-relaxation switching (applied only to class S), and the disease-induced natural mortality switching (applied only classes I and Ir) functions obtained from MLE fits of the relevant parameters to the first 100 days of data (see Fig. 7) are shown over the interval March 1 to August 18. The onset values for these three switching functions are days 4 (March 5), 74 (May 14), and 0 (March 1) respectively. The switching points (half way between initial and final values, although the switching time can precede the onset time, as discussed in Section A.3 and caption to Fig. A.2, SM) are on days 6 (March 7), 38 (April 8) and 4 (March 3) respectively. The range of initial to final values are indicated by the values that scale the vertical axes of the three curves in question.

Phase 4 of the Israeli COVID-19 outbreak, starting around early to mid-July, exhibited considerably more oscillations in daily incidence and less directionality in growth (phases 1 and 3) or decline (phase 2) associated with it (Fig. 4) than the first three phases. Thus we speculate that phase 4 represents a mixture of different intensities of social distancing and social relaxation at different times in different parts of Israel, as various categories of workers and groups of individuals change behaviour in response to social policies and individuals’ perceptions of risk. Thus, we decided to accept the MLE parameters up to day 125 (July 4), as fitted to the first 135 days of incidence and mortality data (Fig. 6) and to carryout MCMC fits of the incidence data from day 128 to 166 (as shown in Fig. 9) with respect to constant social distancing and relaxation rates, as described in Appendix C.3 (SM). More precisely, we carried out an MCMC fit of `SocDist_const` and `SocRel_const` (cf. bottom row of sliders in Fig. 3) on the incidence data interval [125,166], with all other parameters fixed at the values associated with the fits depicted in Fig. 7.

**Figure 9.**
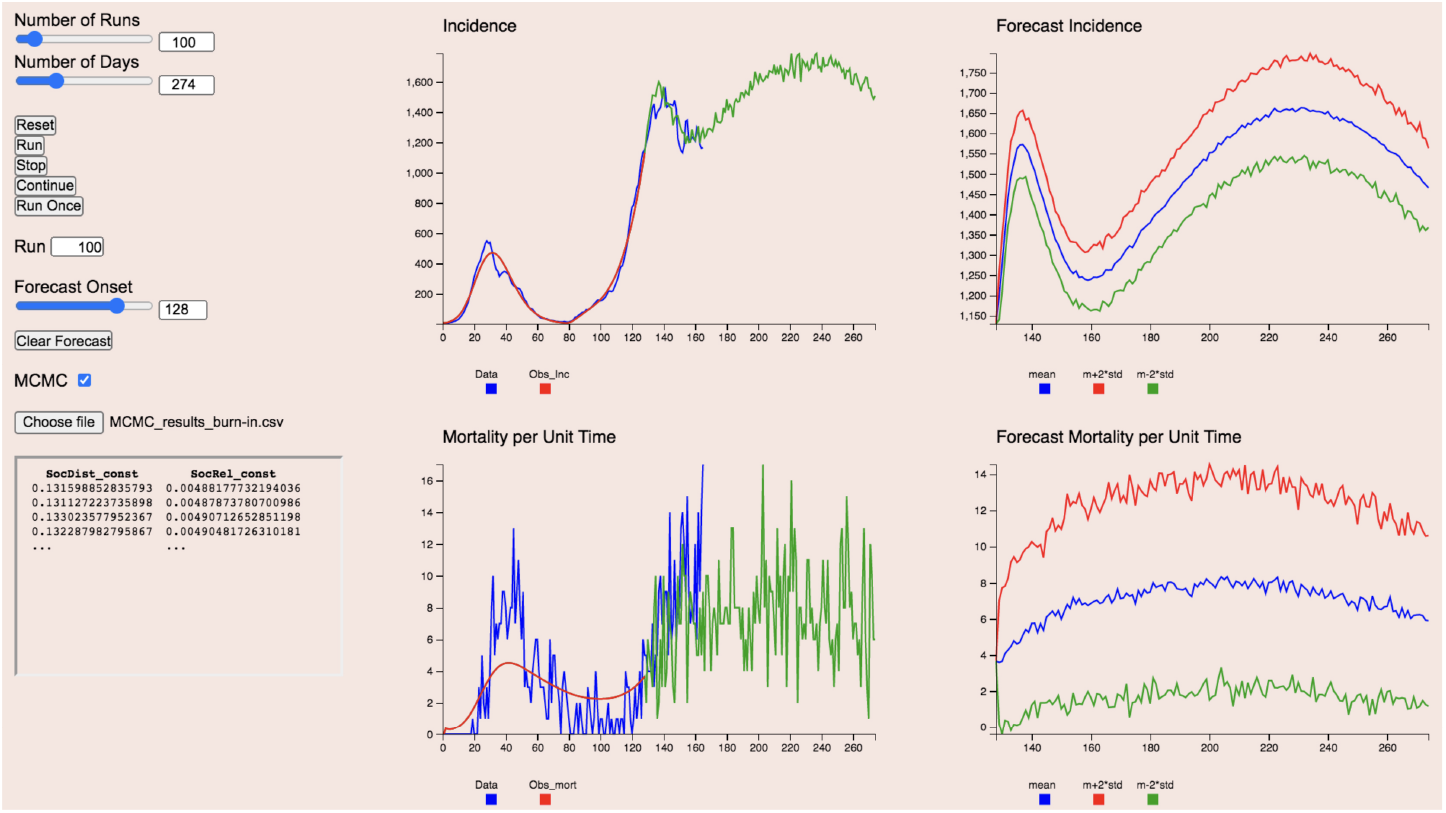
A partial view of the Forecasting page show the Number of Runs to be performed, The number of Days for the total simulations, the Forecast Onset day and the MCMC box checked with the first four rows of MCMC input data shown. The first part of the model projections of incidence and mortality per unit time are in red in the two left most graphics panels, along with the data plotted in blue. The green curves beyond the red are single stochastic simulations (since the Deter Stoc switch is flipped to the Stoc). The two right side panels represent the mean plus/minus 2 standard deviations generated from 100 runs using the MCMC data and stochastic projection mode. The number of runs made is specified in the Number of Runs window.

The distribution of the parameters obtained from these MCMC fits are provided in Appendix C (SM). The actual forecasts are illustrated in Fig. 9. The plotted standard deviations are based on 100 runs. The mean values `SocDist_const`=0.1316 and `SocRel_const`=0.00487 obtained from the MCMC procedure respectively represent 103% and 27% of the values of the social-distancing and social-relaxation switching functions on day *t* = 125 (Fig. 8), which are: `SocDist`(125)=0.127 and `SocRel`(125)=0.018. The MCMC estimates of these two constants are highly correlated (the cross correlation is 0.984; Appendix C.3, SM). Thus, essentially, it appears that a four-fold reduction in the social-distancing levels at the beginning of July have confined incidence to the relatively high levels of 1100 to 1700 cases per day throughout September to November. It will require an additional 5-fold reduction in the social distancing level of early July to bring the epidemic down the point where the number of new cases per day September to November is below 200 (see Fig. D.2 in Appendix D, SOM, for deterministic projects that verify these ballpark figures).

### COVID-19 in South Africa

The first case of COVID-19 was recorded in South Africa on March 5. A week later only 1 of the 17 recorded cases appeared to involve a local transmission event ([VOA News, March 12, 2020](https://www.voanews.com/science-health/coronavirus-outbreak/south-africa-sees-first-local-coronavirus-transmission)): most new cases during the first three weeks of March 2019 were associated with individuals returning from trips to Europe. The local outbreak only started to pick up the last week of March. This local importation effect is evident in the new cases plotted in Fig 10A (see region around the last week of March). Thus we decided to only start fitting the incidence data from Apri 1 onwards. As with the Israeli study, we fitted our model to a 7-day lagged moving averages (Fig 10A and B), rather than the raw data themselves.

**Figure 10.**
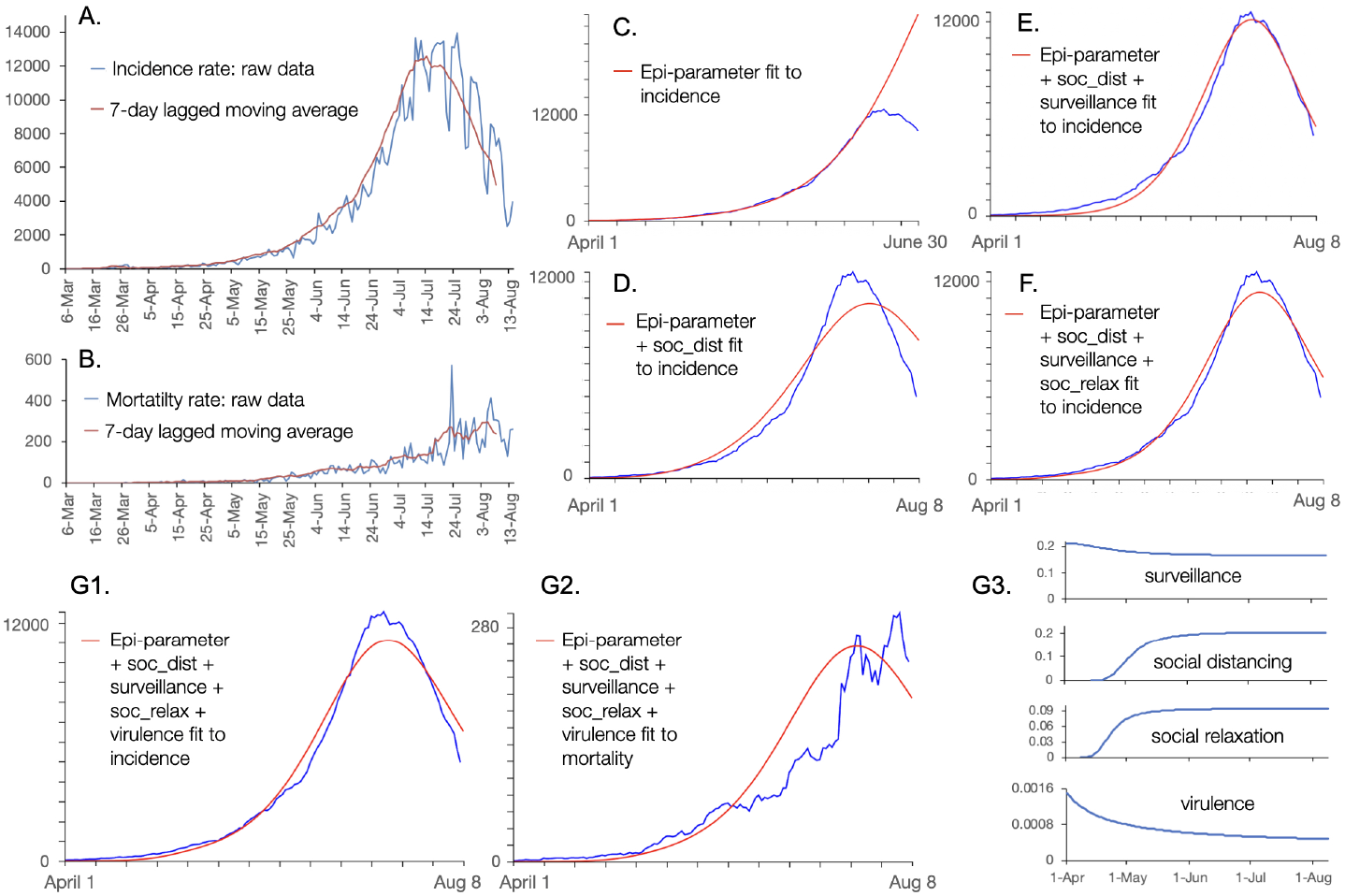
A. The incidence (daily new cases; blue) data time series and a 7-day lagged moving average (brown) plotted for new cases of COVID-19 in South Africa from March 6 to Aug 8 (raw data to Aug 14), 2020. B. As in A but plots for daily COVID-19 deaths rather than incidence. C. Maximum-likelihood estimation (MLE) fits (red plot) of the 4 basic SCLAIV parameters (i.e., kappa, Psuc, I0, C0) to the 7-day lagged moving average incidence time series (blue) from April 1 to June 30 (91 points). D.-F. As in C but fitting over the interval April 1 to August 8 and, in addition to 4 basic SCLAIV parameters, progressively including: D. 4 social distancing switching function parameters (SocDist steep fixed at 3); E. 3 surveillance switching function parameters (onset time, Surveil on, fixed at 0; Surveil steep, fixed at 2); and, F. 4 social relaxation switching function parameters (SocRel steep fixed at 3). G. A combined MLE fit of the incidence (G1.) and daily mortality (G2.) time series from April 1 to Aug 8 of the 4 basic SCLAIV parameters plus social distancing, surveillance, social relaxation and switching function parameters, with the MLE forms of these latter four functions presented in plots G3. See Fig. D.6 in Appendix D, SM, for switching parameter values. Incidence and mortality data source: [Our World in Data](https://ourworldindata.org/coronavirus/country/south-africa?country=~ZAF).

Using the step-wise approach described in Section 2.3 and illustrated more fully in the Israeli case study, we conducted a four basic SCLAIV parameter MLE fit of the incidence data from April 1 to June 30 (91 points) and projected 4 weeks beyond this to provide convincing visual evidence (Fig. 10C) that social distancing had started to play a key role in flattening the incidence curve some weeks before. We thus conducted a 4-SCLAIV plus 4 social-distancing parameters MLE fit of the first 130 incidence data points, obtaining the relatively poor fit depicted in (Fig. 10D). This fit dramatically improved when 3 surveillance parameters were added to the MLE optimisation (Fig. 10E). An additional improvement was obtained when including 4 social-relaxation parameters in the MLE optimisation (Fig. 10F). Although Fig. 10E looks like a better fit than Fig. 10F it is not in terms of MLE (the errors associated the fits depicted in Figs. 10E and F respectively are 15643 and 5453: see Figs. D.4 & D.5 in SOM) because poorer fits in the early stages, when cases are few, are penalised more than poor fits in the later stages when cases are many. The reason for this is that the value of log-likelihood function (Eq. C.2 in Appendix C, SOM) is dependent on proportional rather than absolute deviations because computations are based on the Poisson probability of particular values arising. Also, although social relaxation switches on earlier than social distancing, this early onset has no effect until individuals begin to accumulate in the social distancing class S_r_.

### COVID-19 in England

The first two confirmed cases of COVID-19 in England were identified on January 29 in the city of York (Metro News, April 19, 2020). During the month of March another 56 cases in England were recorded ([UK Government Sources](https://coronavirus.data.gov.uk/)). Our analysis starts at this point in fitting our model to the 7-day lagged moving average of new case in England, starting on March 1 and ending on April 30 (Fig. 11C, D & E) or August 10 (Fig. 11F & G).

**Figure 11.**
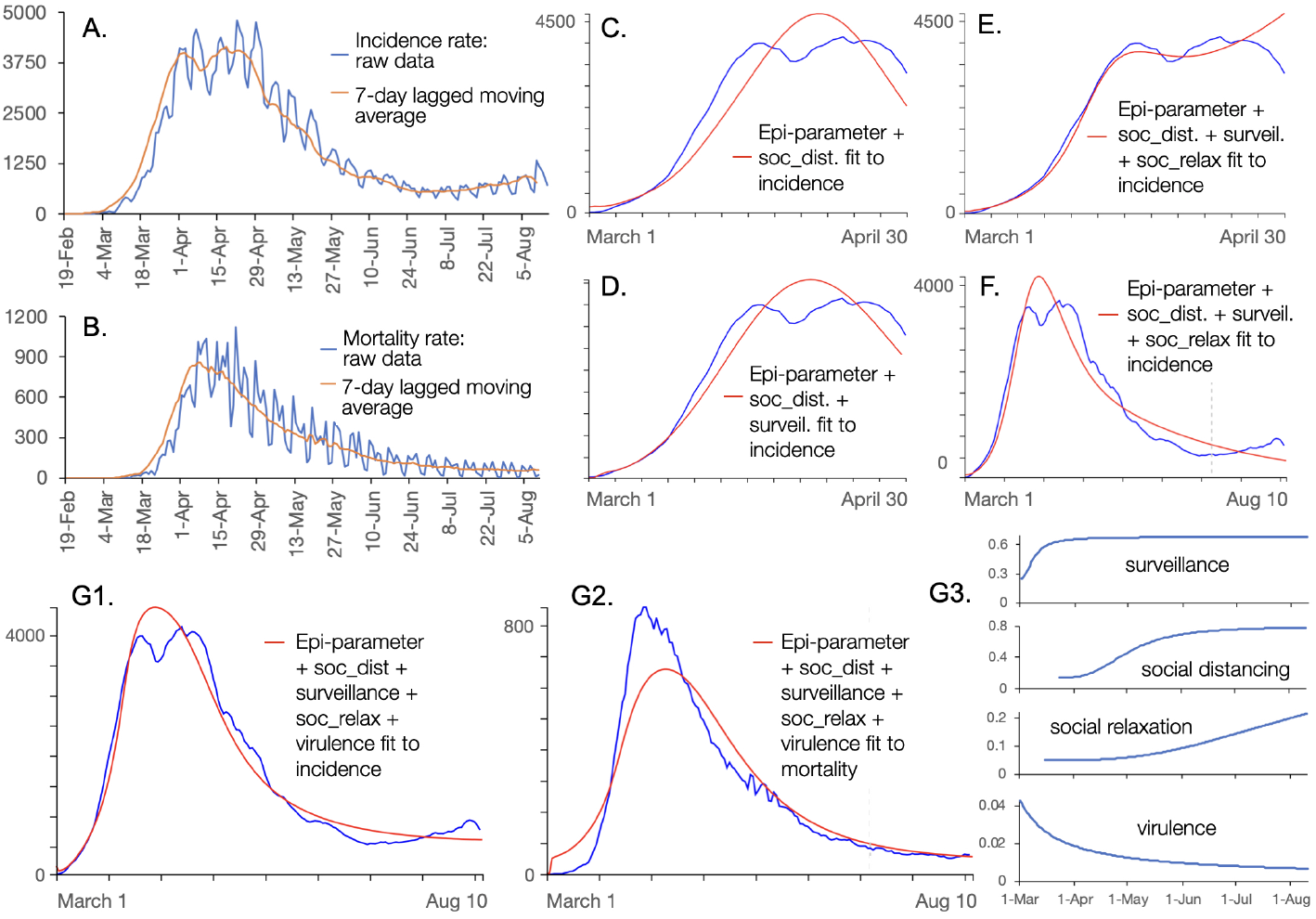
A. The incidence (daily new cases; blue) data time series and a 7-day lagged moving average (brown) plotted for new cases of COVID-19 in England from Feb 19 to Aug 10 (raw data to Aug 16), 2020. B. As in A but plots for daily COVID-19 deaths rather than incidence. C. Maximum-likelihood estimation (MLE) fits (red plot) of the 4 basic SCLAIV parameters (i.e., kappa, Psuc, I_0, C_0) to the 7-day lagged moving average incidence time series (blue) from March 1 to April 30 (first 60 points). D. & E. As in C but in addition to 4 basic SCLAIV parameters, progressively including: D. 4 social distancing and 3 surveillance switching function parameters (SocDist steep, Surveil steep, and Surveil on fixed at 3, 2, and 0 respectively) and E. 4 social relaxation switching function parameters (SocRel steep fixed at 3). F. As in E, but now fitting over the full 163 incidence points. F. G. A combined MLE fit of the incidence (G1.) and daily mortality (G2.) time series from April 1 to Aug 10 of the 4 basic SCLAIV parameters plus social distancing, surveillance, social relaxation and switching function parameters, with the MLE forms of these latter four functions presented in plots G3. See Fig. D.11 in Appendix D, SM, for switching parameter values. Data source: [UK Government Sources](https://coronavirus.data.gov.uk/).

Our first fit of our model to COVID-19 incidence in England over the period March 1 to April 30 involved MLEs of the 4-SCLAIV plus 4 social-distancing switching function parameters. This reasonably fit was notably improved (the absolute loglikelhood error value dropping from 3360 to 1812) by adding 3 surveillance switching function parameters (Fig. 11D). Further improvement was obtained (log-likelhood error value now dropping to 831) by adding 4 social-relaxation switching function parameters to the MLE fit. This 15-parameter MLE fit was then extended to the full 163 point time series (i.e., up to August 10) to obtain the fit depicted in Fig. 11F (for more details on Figs. 11C-F see Figures D.7-10, SM). Rather than accepting this fit to incidence, we carried out simultaneous fit of incidence and mortality with the inclusion of the virulence switching function parameters and setting `Wt:Incidence/Deaths`=0.5. Specifically, we added 3 virulence switching parameters to the 15 used to fit the incidence alone in Fig. 11F to obtain the fit to incidence depicted in Fig. 11G1 and the fit to mortality depicted in Fig. 11G2. The forms of the switching curves obtained from this MLE fit are provided in Fig. 11G3 (see Fig. D.10, SM, for more details and parameter values).

The switching functions depicted in Fig. 11G3 suggest the following. First, surveillance started out around 0.3, but rapidly doubled during the first month (March) to the asymptotic value of 0.6. Social distancing appears to only have started in earnest around the end of March raising four-fold in value through April and May. Social relaxation also started in March (the actual rate has no effect until individuals start to accumulate in the risk reduction class S_r_ through the action of social distancing), rising steadily in through May, June and July as a partial counter to the effects of the social distancing process. This social relaxation counter to social distancing remained below a quarter of the per-capita social distancing level (percapita S for social distancing, per-capita S_r_ for social relaxation) through most of this time, which allowed social distancing to dominate and keep the epidemic decline going throughout all of May and June. In July, however, the effect of social relaxation reversed this trend, and it remains to be seen which will dominate from mid August onwards as social behaviour adapts over time.

## Discussion

The three epidemics that we examined represents a range that includes: one of the most serious epidemics worldwide, which in late August appeared to be well passed its peak (South Africa); one of the most severe epidemics in Europe that now has been brought under control, but could well breakout again if vigilance is lost (England); and a relatively large outbreak for a smaller country that exhibited a second wave considerably more severe than first, and that is now heading into a phase where the epidemic could well fester for the rest of 2020 (Israel). In all of these, our analyses uncovered putative periods of time when different driving forces were dominating: periods when social distancing provided the necessary driving force to flatten and even reverse the exponential growth phase, followed by periods when relaxation of social distancing threatened or allowed a second wave, an including periods of change in surveillance or treatment efficacy. Although statistical modelling is required to link values for these drivers to various activities, their quantification in the context of specific cases allows relative assessments to be made for each of those cases. Thus, we are able to make statements such as: the Israeli epidemic will continue to produce an average of around 1500 new case per day throughout August to November, unless the efficacy of social distancing measures rise to at least 3/4 of where they were throughout March.

Most current models used to study COVID-19 epidemics do not have the kinds of structures in place to facilitate analyses of the spectrum of drivers considered here. Further, web apps available for aiding such analyses are rather limited. The most flexible of these is the [COVID-19 Simulator](https://www.covid19sim.org/), developed by a consortium that includes the Massachusetts General Hospital, Harvard Medical School, Georgia Tech, and Boston Medical Center. The COVID-19 Simulator provides “… a tool designed to help policymakers decide how to respond to the novel coronavirus.” The tool can be used to explore the impact of social-distancing interventions by selecting one of four intensity levels applied at a selected time. The simulator does not allow the user to upload his or her own data and limits analyses to US National and State levels. Other web sites, such as [IDM’s](https://www.idmod.org/documentation) (Institute of Diseas Modeling) EMOD or Covasim, or the platform [EpiModel](https://www.epimodel.org/index.html) require users to have some coding or epidemiological modelling experience.

Strengths of the NMB-DASA web app are that it allows users to input their own data, fit model parameters to data using maximum-likelihood methods, and undertake scenario and forecasting studies with the ability to manipulate an array (8 in total; see Fig. 1) of policy-relevant drivers. Additionally, the South African, English and Israeli COVID-19 case studies provide clear exemplars on how the NMB-DASA Web App can be used to identify which drivers dominate during different phases of an epidemic. If other healthcare and social media infrastructural data (i.e., beyond incidence and mortality rate time series) can be used to independently specify or estimate surveillance [27, 28], social distancing and relaxation [29], quarantining [4] and treatment advances, then further analyses can be undertaken to verify results. In particular, proportions of individuals in the immune class predicted by our model can be tested if appropriately collected serology data are available [30].

Although our exemplar indicates that NMB-DASA can be used to perform analyses of disease outbreaks caused by directly transmissible pathogens for which incidence data alone are available, much deeper analyses can be undertaken. First, spatial structure can be taken into account [21, 31, 32, 33]. Second, links between model output and data may be sought that pertain to individual and societal factors influencing transmission. This can be done by making statistical links between model parameters putative factors [34, 35]. This would require the dynamic analyses of the types demonstrated here augmented with statistical models that linked dynamic measures (such as new cases growth rates over initial and later phases of the outbreak) to key factors of interest.

Critics of our SCLAIV+D+response approach to modeling COVID-19 could say that: 1.) our model has too many parameters for fits to be credible; 2.) the focus is on point estimates to obtain fits of the model to incidence and mortality data, since MCMC fits are currently external to the app; 3.) many of these point estimates are either not well known or possibly not accurate; or, 4.) the results we obtain have not been adequately validated or even their robustness confirmed using sensitivity analysis [36] or other assessment techniques.

In addressing these criticisms, we note Einstein’s dictum that “Everything should be made as simple as possible, but not simpler” speaks to two ideas: the idea of Occam’s razor that the most simple of all formulations needed to capture a particular process should be used; and the idea that if a model is too simple it omits a key process altogether. SEIR models themselves conform to Occam’s razor in ignoring heterogeneities in the relative succeptibility and infectivity of individuals, and in invoking the assumption of a well-mixed population that obviates the need for considerations of spatial structure. Heterogeneity in infectivity is of importance when considering the questions of, say, the role of superspreaders [37], susceptibility and morbidity with regard to age [38], or policies related to particular subgroups in the population, such as opening schools [39]. Spatial structure is important, for example, when considering the impact of restricting travel [40] on managing COVID-19 epidemics. Just as with any SEIR model, the web app can be used to focus on subgroups or spatial structure, although assumptions about links between these groups and the population at large will need to be made.

Second, since the purpose of our web app is to provide a tool for shedding light on the aggregated effects of several different behavioural and mitigating drivers as they ramp up or down over time, we need to include all drivers of interest in the context of making the model adequate to address the questions at hand [41]. In addition, we cannot identify when these drivers may be ramping up or down if we do not characterise them as switching functions. This requires a proliferation of parameters, but these parameters are not all involved in fitting the model at various stages of the outbreak. This is the reason for trying to fit the basic SCLAIV parameters that cannot be independently obtained to the initial outbreak phase and then identifying other phases where the parameters of particular drivers can be estimated. It is also why it is useful (though not necessary) to be able to fit changes in virulence to the mortality data alone once fits to the incidence data have taken place.

Third, not all parameters need to be accurately known (though the more accurate the better), if some are to be specified and others fitted to the data. For example, the initial phases of an epidemic are characterised by its growth rate per unit serial interval time. This characterising value is represented by the quantity *R*_0_, which we defined in the introduction as the average number of individuals an infected individual can expect to infect at the initial stage of an outbreak. Depending on the complexity of the model, estimation of *R*_0_ involves contact rates and infectiousness (force of infection), periods of time spent in disease classes, and even disease-induced mortality rates [42]. Thus, if one of these parameters is not accurately known, our experience has been [33] that the estimation process will compensate by adjusting the values of other parameters involved in the computation of *R*_0_. This compensatory process can be seen quite clearly in the context of the MCMC forecast undertaken for the fourth phase of the Israel outbreak, where we see that that correlation between the social distancing and social relaxation constants being estimated exceeds 98% (Fig. C.2, Appendix C, SM). Additionally, the question of when and where to carry out Bayesian MCMC rather than MLE model fitting of parameters is one that requires a much more extensive discussion than can be undertaken here. We will skirt around this by saying that MCMC is really important when it comes to using forecasts to mitigate risk—that is, when it is important to have a sense of how bad things might get. Maximum likelihood estimation, however, is much easier and faster to implement and, because of its relative simplicity, may be more useful when trying to identify different phases of an outbreak and making comparative assessments on the efficacy of implementing particular mitigating drivers.

Fourth, we agree that sensitivity analyses are useful for evaluating the robustness of results and should be undertaken in studies that probe more deeply than presented here. Additionally, our SCLAIV model provides a number of auxiliary predictions, such as the proportion of individuals that are immune (i.e., the value of the variable *V* (*t*)*/N* (*t*) as it changes over time) that can be evaluated against serological data obtained through a statistically valid sampling method [43]. Deeper in-country studies, involving local experts and a wide array of auxiliary data (i.e., beyond incidence and mortality rate time series) can thus also be used to either further support or refute results obtained.

In summary, we believe our website is based on a model that is both relevant and appropriate [44] for carrying out the types of forensic analyses on social and mitigating drivers of epidemics illustrated in our three COVID-19 case studies and, thereby, provides another tool in the epidemiologist tool box for managing outbreaks. COVID-19 itself is not an anomaly, but one event in series of expected future events that have a past history in SARS, MERS, Ebola, not to mention avian influenza and other highly contagious emerging infectious diseases of zoonotic origin [45, 46, 47]. Our current experience with COVID-19 will help facilitate our response to these future outbreaks and potential pandemics. The availability of computational tools, such as NMB-DASA, will enable policy makers and health-care administrators to be caught less flat-footed at the start of the next outbreak than has been the case for the COVID-19 pandemic.

## Data Availability

This paper scraps data from the web. All sources of data are fully referenced

http://covid-webapp.numerusinc.com/

## Author’s contributions

Wayne M. Getz, conceived the study, formulated the model and wrote the manuscript. Richard Salter developed the web site and the Numerus Modeling Builder software platform. Richard Salter and Ludovica Luisa Vissat were responsible for the MCMC simulations and the material on this in the SM. Nir Horvitz scrapped data from the web. All authors read and edited the final version of the paper.

## Acknowledgements

This research was funded by NSF Grant 2032264.

## Additional Files

### Supplementary Material

A supplementary file is provided containing: Appendix A, details of a continuous-time version of our SCLAIV model; Appendix B, our discrete-time implementation of this model; Appendix C, parameter estimation and simulation procedures, including details of our MCMC computation; and Appendix D, additional case-study figures.

### Training Videos

Three training videos are available with links provided by selecting the instructional material button at the NMB-DASA Home page. Direct links to these videos are: Training video 1, [Introduction to the Numerus Data and Simulation Analyses (DASA) Website for Covid-19](https://www.youtube.com/watch?v=iK4VSdiMOC4); Training video 2, [Parameter Estimation Through Model Fitting](https://www.youtube.com/watch?v=ZBdwZaknyj4&t=604s); Traning video 3, [Deterministic and Stochastic Forecasting](https://www.youtube.com/watch?v=NtwcCGDog_o)

